# Adiposity, metabolites and endometrial cancer risk: Mendelian randomization and Observational analyses

**DOI:** 10.1101/2024.04.18.24305987

**Authors:** Matthew A Lee, Vanessa Y. Tan, Dimitri J Pournaras, Sabrina Wang, Laure Dossus, Marc J. Gunter, Kaitlin H. Wade, Laura J. Corbin, Nicholas J. Timpson

**Author notes:** **Corresponding author:** Professor Nicholas J. Timpson, Address: MRC Integrative Epidemiology Unit, University of Bristol, Oakfield House, Oakfield Grove, Bristol, BS8 2BN. These authors contributed equally to this work (Matthew A Lee, Vanessa Y. Tan).

## Abstract

**Introduction:** Increased and excess adiposity is associated with increased risk of endometrial cancer (EC) and both of these are associated with circulating metabolite profiles. However, how metabolites relate to the adiposity-EC relationship remains unclear.

**Methods:** We have brought together evidence from Mendelian randomization (MR) and observational analyses to evaluate the effect of i) adiposity traits on endometrial cancer, ii) adiposity traits on circulating metabolites and iii) adiposity-associated metabolites on EC. We have also evaluated the potential role of metabolites in the adiposity-EC relationship using multivariable MR. Observational analyses were conducted using individual level data from UK Biobank (N = 1,005 cases and 215,339 controls). MR analyses were performed using female-specific summary statistics from genome-wide association studies of body mass index (BMI; N up to 434,794), waist-to-hip ratio (WHR; N up to 381,152), 249 metabolites and ratios from targeted nuclear magnetic resonance metabolomics (N up to 140,768) and EC risk (12,906 cases and 108,979 controls).

**Results:** In observational analyses, higher BMI and WHR were associated with elevated odds of overall EC (odds ratio (OR) per standard deviation (SD) increase in BMI = 1.37; 95% confidence interval (CI) = 1.19, 1.57; OR per SD increase in WHR= 1.15; 95% CI = 1.01, 1.32). In MR analysis, higher BMI was associated with elevated odds of overall EC risk (OR per SD increase in BMI = 1.80; 95% CI = 1.56, 2.07), endometrioid cancer (OR = 1.71; 95% CI = 1.45, 2.02) and non-endometrioid cancer (OR = 2.20; 95% CI = 1.55, 3.12). There was weaker evidence for a causal relationship with WHR. BMI was associated with 165 metabolites and ratios after Bonferroni-correction in MR analyses, several of which were associated with EC and 25 of which were directionally consistent with an intermediate role in the effect of BMI on EC risk from two-step MR and observational analyses. In MVMR analyses, there was evidence suggesting that the effect of BMI on non-endometrioid EC was mediated by several lipid metabolites; for example, the univariable MR OR for non-endometrioid EC per 1 SD increase in BMI was 2.51 (95%CI = 1.47, 4.29), whereas on adjusting for free cholesterol to total lipids ratio in medium LDL, the MVMR OR for non-endometrioid EC per 1 SD increase in BMI was 1.18 (95%CI = 0.53, 2.66). Further bioinformatic analyses highlighted a mixture of other potential shared pathways (including height, adiposity traits and blood cell traits) that could influence the risk of EC.

**Conclusion:** Evidence here suggests that higher BMI causes a higher risk of overall and all histological subtypes of EC and variation in numerous circulating metabolites. Several of these metabolites showed relationships consistent with an intermediate role between BMI and non-endometrioid EC, however, further bioinformatic analyses highlighted other potential shared mechanisms that could influence the risk of EC.

## Introduction

Endometrial cancer (EC; cancer of the lining of the uterus) is the most common gynaecological cancer among women with more than 380,000 new cases diagnosed globally^1^. Based on differences in histology and clinical outcomes, there are two main subtypes of EC: endometrioid carcinomas (type 1 tumours) with good prognosis and non-endometrioid carcinomas (type 2 tumours) with worse prognosis^2^. Endometrioid EC is more commonly hormonally driven compared with non-endometrioid EC^3^.

Excess body weight is robustly associated with EC, with support from observational studies^4^. Mendelian randomisation^5^ (MR) studies, which use genetic variants as instruments (or proxies), of adiposity traits further support the evidence base for a causal relationship with EC^6–8^. However, whilst the relationship between adiposity and EC is well-established, understanding of the mechanisms and covariables involved in the adiposity-EC relationship remains incomplete. Furthermore, evidence of associations between body fat distribution, measured as waist-hip-ratio (WHR), has only been supported with observational studies^4,6,8^.

Metabolic reprogramming is recognised as a hallmark of tumorigenesis^9^ and there is evidence that metabolic dysfunction drives the development and progression of EC^10,11^. Findings from a recent prospective study suggest that concentrations of glycine, serine, sphingomyelin and free carnitine may represent specific pathways involved in EC development^12^. Increased adiposity causes changes to an individual’s systemic metabolic profile^13–15^. Observational and MR studies support an effect of adiposity (proxied using body mass index [BMI]), on raised amino acids, fatty acids and inflammatory glycoprotein acetyls^15^, leading to suggestions that a potential mechanism linking adiposity and EC could be adiposity-induced metabolic changes^10^. A recent prospective study showed that EC is positively associated with adiposity-associated metabolic changes including specific amino acids and lipids^16^; however, whether these relationships are causal is unclear.

We aimed to better understand the potential role of circulating metabolites as intermediates in the association between adiposity and EC risk by triangulating evidence from summary-level univariable two-step^17^ MR and multivariable MR approaches in combination with observational analyses in UK Biobank.

## Methods

### Analytical strategy

This study has four main analyses that were performed sequentially (**Figure 1**) to estimate: (**Part I**) the effect of adiposity measures on EC, (**Part II**) the effect of adiposity measures on circulating metabolites, (**Part III**) the effect of adiposity-associated metabolites on EC, and (**Part IV**) the potential intermediate role of adiposity-associated metabolites in the relationship between adiposity and EC (identified in **Part II** and **III**). Observational analyses were performed for Parts I-III. MR analyses were performed for Parts I-IV; Metabolite data from UK Biobank were used in both the MR and observational analyses. This study is reported as per the Strengthening the Reporting of Observational Studies in Epidemiology (STROBE) and STROBE-MR guidelines (**Supplementary Table S1** and **S2**)^18,19^.

**Figure 1.**
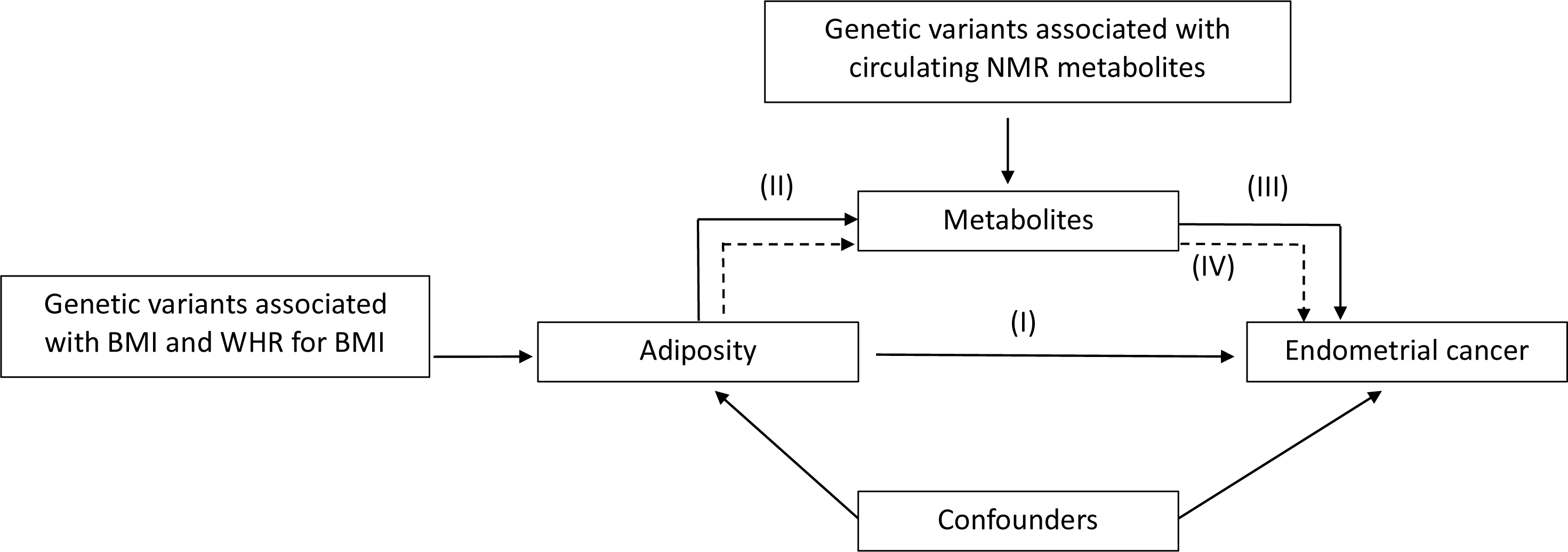
**Study overview.** This study has four main analyses that were performed sequentially to estimate: (**Part I**) the effect of adiposity measures on EC, (**Part II**) the effect of adiposity measures on metabolites, (**Part III**) the effect of adiposity-associated metabolites and EC, and (**Part IV**) the potential intermediate role of adiposity-associated metabolites in the relationship between adiposity and EC (identified in **Part II** and **III**). Conventional observational analyses were performed for Part I-III; Mendelian randomization analyses were performed for Parts I-IV.

### Observational analyses (Part I-III)

#### Study population

UK Biobank is a nation-based health project that recruited more than 500,000 participants (aged 37-73 years, 56.3% were women) between 2006 and 2020^20,21^. In addition to the collection of biological samples (blood, saliva and urine), health, demographic and anthropometric data were collected in 22 assessment facilities across England, Wales and Scotland^22^. Participants provided written informed consent. Ethical approval was obtained from the Northwest Multi-centre Research Ethics Committee (11/NW/0382). Data from UK Biobank were accessed via application number 16391 and 30418. Data for adiposity measures and endometrial cancer outcome was extracted on the 08/07/2021 and the ^1^H-NMR metabolite data was extracted on the 08/02/2023.

##### Adiposity measures

BMI was calculated as weight (kg)/height^2^ (m^2^) and WHR as waist circumference (cm)/hip circumference (cm). Height was measured to the nearest centimetre, using a Seca 202 stadiometer, and body weight to the nearest 0.1kg, using a Tanita BC-418 body composition analyser. Waist circumference was measured at the natural indent (the umbilicus was used if the natural indent could not be observed); hip circumference was measured at the widest part of the hips. BMI and WHR were inverse rank normal transformed prior to observational analyses and represent normalised SD units.

##### Metabolite measures

Non-fasting EDTA plasma samples were collected from approximately 275,000 participants, a random subset of the original ∼500,000, who provided samples at the 2006-2010 assessment (N=275,000) or at a subsequent assessment in 2012-2013 (N=17,000) (around 15,000 of these have both baseline and repeat assessment). A total of 249 metabolic traits (168 concentrations plus 81 ratios) were quantified using ^1^H-NMR spectroscopy (data pre-processing and quality control steps conducted by Nightingale Health are described previously^23,24^). Inverse rank normal transformed metabolite concentrations were used in all analyses and represent normalised SD units.

##### Endometrial cancer

EC was defined as a malignant neoplasm of the endometrium. The UK Biobank database contained a record of all cancers including their subtype occurring either before or after participant enrolment using the International Classification of Diseases, 9^th^ and 10^th^ revision (ICD-10, ICD-9). The following ICD-10 and ICD-9 codes were used to define EC: ICD10 codes (C540, C541, C542, C543, C549 and C55) and ICD9 codes (179, 1799, 180, 182, 1820, 1821 and 1828). Cases were characterised as incident or prevalent using ‘age when they attended the centre’ and ‘age when first reported EC cancer’. Participants were defined as incident cases if their ‘attending age’ was less than their ‘cancer diagnosis age’. In total, there were 1,935 EC cases with 1,005 being incident cases and 930 prevalent cases. Only incident cases (N=1,005) were included in our analyses. Controls (n=215,339) were defined as female participants who had no record of any type of cancer, in-situ carcinoma, or an undefined neoplasm.

##### Covariables

We included potential confounders of the BMI-metabolite-EC relationships in our analyses. The potential confounders were sociodemographic factors (age at assessment, physical activity, smoking status, alcohol consumption and educational attainment) and female-specific factors (history of hormone replacement therapy (HRT) use, age at first live birth, age at last live birth, age at menarche and menopausal status). Age, smoking status, alcohol consumption, education attainment and female-specific factors were self-reported at the baseline assessment by questionnaire. Physical activity level over a typical week was self-reported using the International Physical Activity Questionnaire and reported as metabolic equivalent of task (MET) per week.

### Statistical analyses

Observational associations between: I) adiposity measures and EC (**Figure 1, Part I**) were assessed using multivariable logistic regression, II) adiposity measures and metabolites (**Figure 1, Part II**) were assessed using multivariable linear regression for cases and controls combined in one cohort and III) adiposity-associated metabolites and EC (**Figure 1, Part III**) were assessed using multivariable logistic regression. All analyses were initially adjusted for age at assessment and centre. Models were additionally adjusted for potential confounders (smoking status, alcohol consumption, education status, physical activity, hormone replacement therapy use, age at first live birth, age at last live birth, age at menarche and menopausal status). To account for multiple testing, we used the Bonferroni-adjusted *p* value threshold (0.05/249) for the association between adiposity measures and metabolites and *p*-value < 0.05 for all other tests. Multivariable linear and logistic regression analyses were performed using the lm and glm functions, respectively, in R.

### Genome-wide association study results and Mendelian randomisation analyses (Part I-IV)

#### Data sources and study populations

##### Adiposity instruments

We identified single nucleotide polymorphisms (SNPs) that were independently associated (low linkage disequilibrium (LD), R^2^<0.001) with BMI and WHR (unadjusted for BMI) at *p*<5x10^-9^ from a recent large-scale female-specific genome-wide association study (GWAS) meta-analysis of 434,794 female adults of European ancestries from the Genetic Investigation of Anthropometric Traits (GIANT) consortium and the UK Biobank^25^ (**Supplementary Table 3**). Adiposity measures were inverse rank normal transformed prior to genome-wide analysis and units therefore represent a normalised standard deviation (SD). In total, 271 (average F-statistic = 41) and 227 (average F-statistic = 45) SNPs were identified for BMI and WHR, respectively.

##### Metabolite GWAS in UK Biobank and selection of metabolite instruments

A random subset of non-fasting baseline plasma samples, consisting of 275,000 UK Biobank participants and 17,000 repeat-visit samples (around 15,500 of these have both a baseline and repeat assessment), were measured using targeted high-throughput ^1^H-Nuclear magnetic resonance (^1^H-NMR) metabolomics (Nightingale Health Ltd; biomarker quantification version 2020)^24,26^. Genotype data was available for 488,377 individuals, of which 49,979 were genotyped using the UK BiLEVE array and 438,398 using the UK Biobank axiom array. Pre-imputation QC, phasing and imputation have been described previously^21^. Genotype imputation was performed using IMPUTE2 algorithms^27^ to a reference set combining the UK 10K haplotype and HRC reference panels^28^. Post-imputation QC was performed as described in the “UK Biobank Genetic Data: MRC-IEU Quality Control” documentation^29,30^. We conducted a female-specific GWAS for the 249 ^1^H-NMR-derived metabolites and ratios in UK Biobank female participants of European descent (N=140,768) using the MRC IEU UK Biobank GWAS pipeline^30^. We restricted the samples to individuals of European ancestry as defined by the largest cluster in an in-house k-means clustering of genetic ancestry data (K=4) after standard exclusions including withdrawn consent, mismatch between genetic and reported sex and putative sex chromosome aneuploidy^29–31^ (**Supplementary Table 3**). Metabolite measures were inverse rank normal transformed prior to genome-wide analysis and units therefore represent a normalised SD. Genome-wide association analysis was conducted using a linear mixed model (LMM) as implemented in BOLT-LMM (v2.3)^32^. Population structure was modelled using 143,006 directly genotyped SNPs (MAF > 0.01; genotyping rate > 0.015; Hardy-Weinberg equilibrium p-value < 0.0001 and LD pruning to an r2 threshold of 0.1 using PLINK(v2.00)). Genotype array and fasting time were adjusted for in the model. We identified 11-167 independent SNPs (r^2^<0.001 and *p*<5x10^-8^) for each of the ^1^H-NMR-derived metabolites (average F-statistics: ranged from 40.6-81.6).

##### Endometrial Cancer GWAS data

We obtained SNP estimates from the largest GWAS (as log odds ratio (OR)) for EC to date^8^, including up to 12,906 cases and 108,979 controls from 13 studies (**Supplementary Table 3**). Of the 12,906 cases and 108,979 controls, 636 cases (5%) and 62,853 controls (58%) were from UK Biobank. Summary statistics were also available for the association between genetic variation and the EC subtypes, endometrioid (8,758 cases) and non-endometrioid (1,230 cases) cancer; both GWASs used the full set of controls (N = 108,979). None of the endometrial subtype cases were from UK Biobank. Histological subtypes of EC were confirmed based on pathology reports and detailed study descriptions have previously been reported^8^.

### Statistical analysis

In each instance, MR estimates are interpreted as the change in outcome per SD unit change in exposure. Estimates for metabolite outcomes reflect SD unit change, and estimates for EC outcomes reflect odds ratios (OR). All analyses were performed using R version 3.5.3^33^. Univariable MR analyses were performed using the TwoSampleMR (version 0.4.22) package^26^. Multivariable MR analyses were performed using the MVMR (version 0.3) package^32^.

First, we examined the association between BMI and WHR with overall, endometrioid and non-endometrioid EC risk using summary-level data from female-specific GWAS of BMI, WHR and EC (**Figure 1, Part I**). Details of the SNPs included in each analysis, and proxies used (where SNPs were not available in the outcome data), are provided in **Supplementary Table 4**. Summary statistics were harmonised using the harmonise_data function within the TwoSampleMR R package (version 0.4.22)^33^. Univariable causal estimates were combined using the inverse-variance weighted (IVW) multiplicative random effects (IVW-MRE) model^34^. Where possible (i.e., where there were three or more instruments), the assumption of no pleiotropy among genetic instruments and outcomes were explored using MR-Egger^35^, weighted median^36^ and weighted mode^37^ estimators. These methods are sensitive to the effects of potential pleiotropy under different assumptions. No *p*-value threshold requirements were set for these methods and, instead, consistency between the IVW model and the three sensitivity MR methods (MR-Egger, weighted median and weighted mode) was assessed.

Second, adiposity traits (BMI and WHR) that showed evidence of an effect on EC with *p* values < 0.05 from IVW-MRE models and with consistent associations across the three sensitivity MR methods, were taken forward and examined for associations with the ^1^H-NMR-measured metabolites (**Figure 1, Part II**). Summary-level data were obtained from a female-specific GWAS for adiposity traits and ^1^H-NMR metabolites, and we examined the effect of adiposity traits on ^1^H-NMR-measured metabolites using the four MR models described above. Metabolites that were associated with the adiposity traits with *p* values < 0.05/249 (Bonferroni-adjusted *p*-value threshold) from IVW-MRE models were taken forward and examined for association with EC risk using IVW-MRE model (if ≥2) or Wald ratio (if 1 SNP) (**Figure 1, Part III**).

Multivariable MR (MVMR) ^38,39^ was conducted to test the hypothesis that adiposity-associated metabolites (identified in **Part II and III**) may act as intermediate factors in the effect of BMI on EC (**Figure 1, Part IV**). Only metabolites that showed a consistent direction of effect across the adiposity-EC, adiposity-metabolite, and metabolite-EC analyses from both MR and observational analyses (identified in **Part II and III**) were included in the MVMR analyses. For the MVMR analyses, we fitted a model with BMI and EC and included each BMI-associated metabolite in turn to estimate the direct causal effect of BMI on EC. We used female-specific SNPs for BMI based on an earlier (lower-powered) GWAS^40^ for the multivariable models to avoid a relative dilution of metabolite instrument strength^41^ given that the number of SNPs for BMI from the latest BMI GWAS far outnumbered those for metabolites. Conditional F-statistics were used to evaluate instrument strength^39^. Heterogeneity was quantified using an adapted version of the Q statistic (Q_A;_ also a further modification of Cochran’s Q)^39^.

### Sensitivity analyses

Sample overlap between exposure and outcome GWAS can bias MR estimates towards the confounded observational estimate (inflated type 1 error) in the presence of weak instrument bias in a manner proportional to the degree of overlap^42^. This bias can be inflated by “Winner’s curse”, in which weights for genetic instruments are derived from discovery samples that overlap with outcome samples. There was sample overlap across our MR analyses as the adiposity, ^1^H-NMR metabolite and EC GWAS all included participants from UK Biobank. Given the random selection of samples for metabolomics analysis and the inclusion of almost all samples within the adiposity GWAS it is not possible to precisely quantify the degree of sample overlap but we make the assumption of 100% sample overlap between the adiposity and ^1^H-NMR metabolite GWAS. There was also approximately 5% overlap for both the adiposity and ^1^H-NMR metabolite GWAS with the overall EC GWAS. Given this, we conducted sensitivity analyses to evaluate the influence of sample overlap in our MR analyses. First, for analyses examining the association between adiposity traits with endometrial cancer risk (**Figure 1, Part I**), we re-performed MR analyses using alternative GWAS data for BMI (n=171,977) and WHR (N=118,004) where there was no sample overlap^40,43^ with either the EC GWAS conducted by ECAC^8^ or an alternative EC GWAS conducted by ECAC which excluded UK Biobank participants (personal correspondence) (N for overall EC=12,270 cases and 46,126 controls; N for endometrioid EC=8,758 cases and 46,126 controls and N for non-endometrioid EC = 1,230 cases and 35,447 controls)^8^. Second, for analyses examining the association between adiposity traits with ^1^H-NMR metabolites (**Figure 1, Part II**), we re-performed MR analyses using alternative GWAS data for BMI where there was no sample overlap^40,43^. Third, for analyses examining the association between ^1^H-NMR metabolites and endometrial cancer risk (**Figure 1, Part III**), we re-performed MR analyses using the alternative GWAS for EC which had excluded UK Biobank participants^8^ (**Supplementary Table 3**).

### Examining off-target effects using PhenoScanner (Part V)

Due to the shared genetic architecture and pleiotropic nature of instruments for metabolites^44–46^, it is possible for variants acting as proxies for metabolites to exert off-target or pleiotropic effects on EC through other biological mechanisms. This presents the possibility that intermediate associations are reflecting common, but unmeasured, biological underpinnings rather than more obvious hypothesised pathway effects. To investigate this, we assessed whether genetic instruments for adiposity- and EC-associated, (and apparently intermediate) metabolites were associated with other traits (*p*<1E-10) in a manner different to that expected by chance given documented genotype/metabolite/trait associations. Using PhenoScanner^47,48^, we assessed the co-association of adiposity- and EC-associated metabolites with other phenotypes and outcomes before performing the same procedure over 100 iterations, but for randomly selected metabolites taken from the 244 metabolites not found to underlie the association between adiposity and EC risk. The overall profile of these redraws were compared with that from the adiposity- and EC-associated instruments, i.e. counts of detectable associations with trait domains for adiposity- and EC-associated instruments versus SNPs associated with random metabolites.

## Results

### Population and data overview

The observational analyses within UK Biobank included up to 1,005 female participants who had a diagnosis of incident EC and up to 215,339 controls (**Table 1**). The cases and controls had a mean (SD) age of 59.63 (6.43) years and 55.83 (8.02) years, respectively. Mean BMI was higher among cases than controls (at 30.33kg/m^2^ (7.07) and 27.06kg/m^2^ (5.17), respectively). Mean WHR was higher among cases than controls (at 0.84 (0.07) and 0.82 (0.07), respectively). 3.66% reported having a diagnosis of diabetes, and this was more common among cases than controls (8.99% and 2.64%, respectively).

**Table 1.** Baseline characteristics of UK Biobank participants included in our study.

|  | N<br>(Cases/Control) | Endometrial cancer<br>Cases<br>(N=1,005) | Controls<br>(N=215,339) |
| --- | --- | --- | --- |
| **Continuous variables** |  |  |  |
| Age at assessment (year) | 1,005/215,339 | 59.63 (6.43) | 55.83 (8.02) |
| Body mass index (kg/m<sup>2</sup>), mean (SD) | 979/211,756 | 30.33 (7.07) | 27.06 (5.17) |
| Waist to Hip Ratio, mean (SD) | 1,000/214,835 | 0.84 (0.07) | 0.82 (0.07) |
| Age at first birth (year) | 632/145,456 | 24.54 (4.23) | 25.38 (4.64) |
| Age at last birth (year) | 632/145,149 | 29.32 (4.55) | 30.35 (4.90) |
| Menarche age (year) | 971/208,424 | 12.64 (1.55) | 12.98 (1.62) |
| **Categorical variables** |  |  |  |
| **Smoking** | 997/214,285 |  |  |
| Never |  | 643 (64.49%) | 129,857 (60.60%) |
| Previous |  | 303 (30.39%) | 65,775 (30.70%) |
| Current |  | 51 (5%) | 18,653 (8.70%) |
| **Alcohol intake** | 1,002/214,865 |  |  |
| Daily or almost daily |  | 143 (14.27%) | 34,152 (15.89%) |
| 3-4 times a week |  | 160 (15.97%) | 44,464 (20.69%) |
| Once or twice a week |  | 252 (25.15%) | 55,476 (25.82%) |
| 1-3 times a month |  | 139 (13.87%) | 28,241 (13.14%) |
| Special occasions only |  | 193 (19.26%) | 32,134 (14.96%) |
| Never |  | 115 (11.48%) | 20,398 (9.49%) |
| **Diabetes** | 1,001/214,512 |  |  |
| Yes |  | 90 (8.99%) | 7,807 (2.64%) |
| No |  | 911 (91%) | 206,705 (96.36%) |
| **Education** | 774/177,857 |  |  |
| College/University degree |  | 304 (39.28%) | 68,385 (38.45%) |
| A levels/AS levels or equivalent |  | 106 (13.70%) | 25,756 (14.48%) |
| O levels/GCSEs or equivalent |  | 215 (27.78%) | 49,943 (28.08%) |
| CSEs or equivalent |  | 49 (6.33%) | 11,924 (6.70%) |
| NVQ or HND or HNC or equivalent |  | 33 (4.26%) | 9,676 (5.44%) |
| Other professional qualifications |  | 67 (8.66%) | 12,173 (6.84%) |
| **Physical activity (No. of days/week of vigorous physical activity)** | 923/202,664 |  |  |
| 0 days |  | 394 (42.69%) | 80,448 (39.70%) |
| 1 days |  | 156 (16.90%) | 29,567 (14.59%) |
| 2 days |  | 148 (16.03%) | 32,675 (16.12%) |
| 3 days |  | 106 (11.48%) | 28,260 (13.94%) |
| 4 days |  | 46 (4.98%) | 12,212 (6.02%) |
| 5 days |  | 36 (3.9%) | 10,652 (5.26%) |
| 6 days |  | 12 (1.3%) | 2,676 (1.32%) |
| 7 days |  | 25 (2.71%) | 6,714 (3.31%) |
| **Had Menopause** | 1,002/214,649 |  |  |
| Yes |  | 822 (82.04%) | 126,379 (58.88%) |
| No |  | 180 (17.96%) | 88,270 (41.12%) |
| **Ever used HRT** | 999/214,185 |  |  |
| Yes |  | 382 (38.24%) | 78,769 (36.78%) |
| No |  | 617 (61.76%) | 135,416 (63.22%) |

kg, kilograms; m<sup>2</sup>, meters squared; SD, standard deviation; p, p value; HRT, hormone replacement therapy; N, sample size

### Association between adiposity measures and endometrial cancer (Part I)

For BMI, observational analyses showed evidence that higher BMI (per SD) was associated with 1.61 (95% CI = 1.49, 1.75) times higher odds of overall EC; these results were consistent when adjusting for all covariables (OR = 1.37; 95% CI = 1.19, 1.57) (**Figure 2**; **Table 2**). This was supported by univariable MR analysis, which found that higher BMI (per SD) was associated with 1.80 (95% confidence interval (CI) = 1.56, 2.07) times higher odds of overall EC using the IVW-MRE model (**Figure 2; Supplementary Table 5**). Similar results were found for endometroid cancer (OR = 1.71; 95% CI = 1.45, 2.02) and non-endometrioid cancer (OR = 2.20; 95% CI = 1.55, 3.12) (**Supplementary Table 5**). This finding was consistent across sensitivity analyses using methods that consider potential genetic pleiotropy and when using instruments from non-overlapping samples (**Supplementary Table 5**).

**Figure 2.**
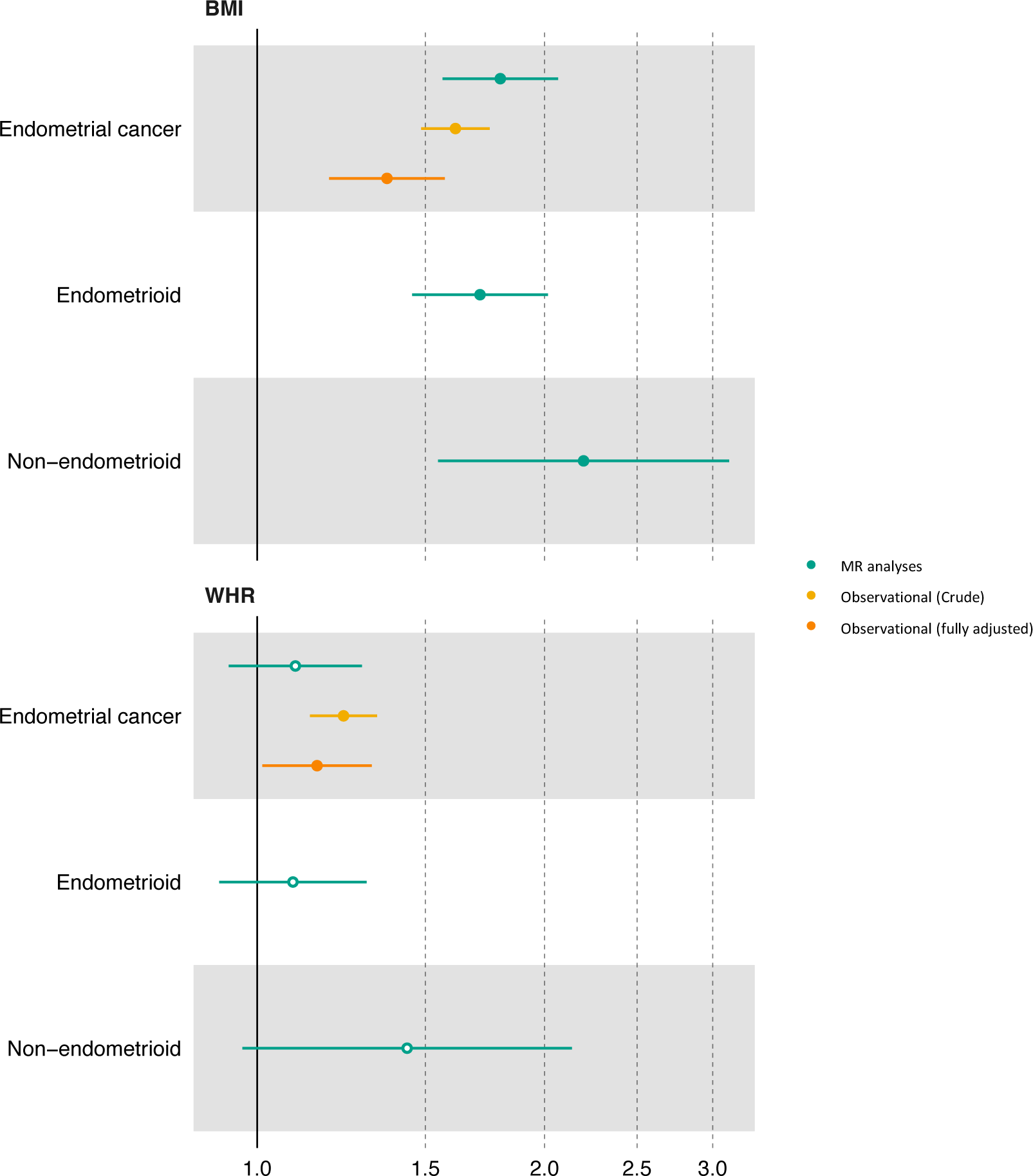
**Mendelian randomization and observational estimates of the effect of BMI and WHR on endometrial cancer** The forest plot shows the estimates of the association between BMI and WHR with overall, endometrioid and non-endometrioid endometrial cancer from MR analyses (IVW method) using summary data from the Endometrial Cancer Association Consortium (ECAC) (n=12,906 endometrial cancer cases and 108,979 controls) and conventional observational analyses using individual level data from UK Biobank. Symbols represent point estimates from individual analyses. Horizontal lines represent 95% confidence intervals. Close circles represent *p*<0.05.

**Table 2.** Observational estimates of the association between adiposity measures and endometrial cancer.

| Exposure | Outcome | Adjusted for age and centre |  | Fully adjusted |  |
| --- | --- | --- | --- | --- | --- |
|  |  | OR<br>(95%CI) | *p* | OR<br>(95%CI) | *p* |
| **BMI** | **Incident EC** | 1.61<br>(1.49, 1.75) | $7.63 \times 10^{-30}$ | 1.37<br>(1.19, 1.57) | $1.07 \times 10^{-5}$ |
| **WHR** | **Incident EC** | 1.23<br>(1.14, 1.34) | $4.30 \times 10^{-07}$ | 1.15<br>(1.01, 1.32) | 0.03 |

BMI, Body mass index; WHR, waist hip ratio; EC, endometrial cancer; OR, odds ratio; CI, confidence interval; *p*, *p* value

For WHR, observational analyses showed evidence that higher WHR (per SD) was associated with 1.23 (95% CI = 1.13, 1.34) times higher odds of overall EC; these results were consistent when adjusting for all covariables (OR = 1.15; 95% CI = 1.01, 1.32) (**Figure 2**; **Table 2**). Evidence of association between WHR (per SD) and overall EC (OR = 1.10; 95% CI = 0.93, 1.29), endometroid EC (OR = 1.09; 95% CI = 0.91, 1.30) and non-endometrioid EC (OR = 1.43; 95% CI = 0.96, 2.12) in univariable MR was weaker (**Figure 2**; **Supplementary Table 5**). Sensitivity analyses were broadly consistent for overall and endometrioid EC. However, findings were less consistent for non-endometrioid EC in sensitivity analyses where the MR-Egger estimate (OR = 3.99; 95% CI = 1.14; 13.95) was greater in magnitude compared to the IVW estimates (**Supplementary Table 5**) and when using instruments from non-overlapping samples (IVW OR= 1.93; 95% CI = 1.17, 3.18) (**Supplementary Table 5**).

### Association between adiposity measures and metabolites (Part II)

Higher BMI (per SD) was associated with 175 metabolites after correcting for multiple testing and using the IVW-MRE model (**Figure 3**; **Supplementary Table 6**). Of these metabolites, 165 had consistent directions of effect in all four MR models and the fully-adjusted observational analyses (**Supplementary Figure 1 and 2**; **Supplementary Table 7)** and were taken forward for subsequent analysis with EC. These findings were broadly consistent across sensitivity analyses using instruments from non-overlapping samples (r=0.94) (**Supplementary Table 6**).

**Figure 3.**
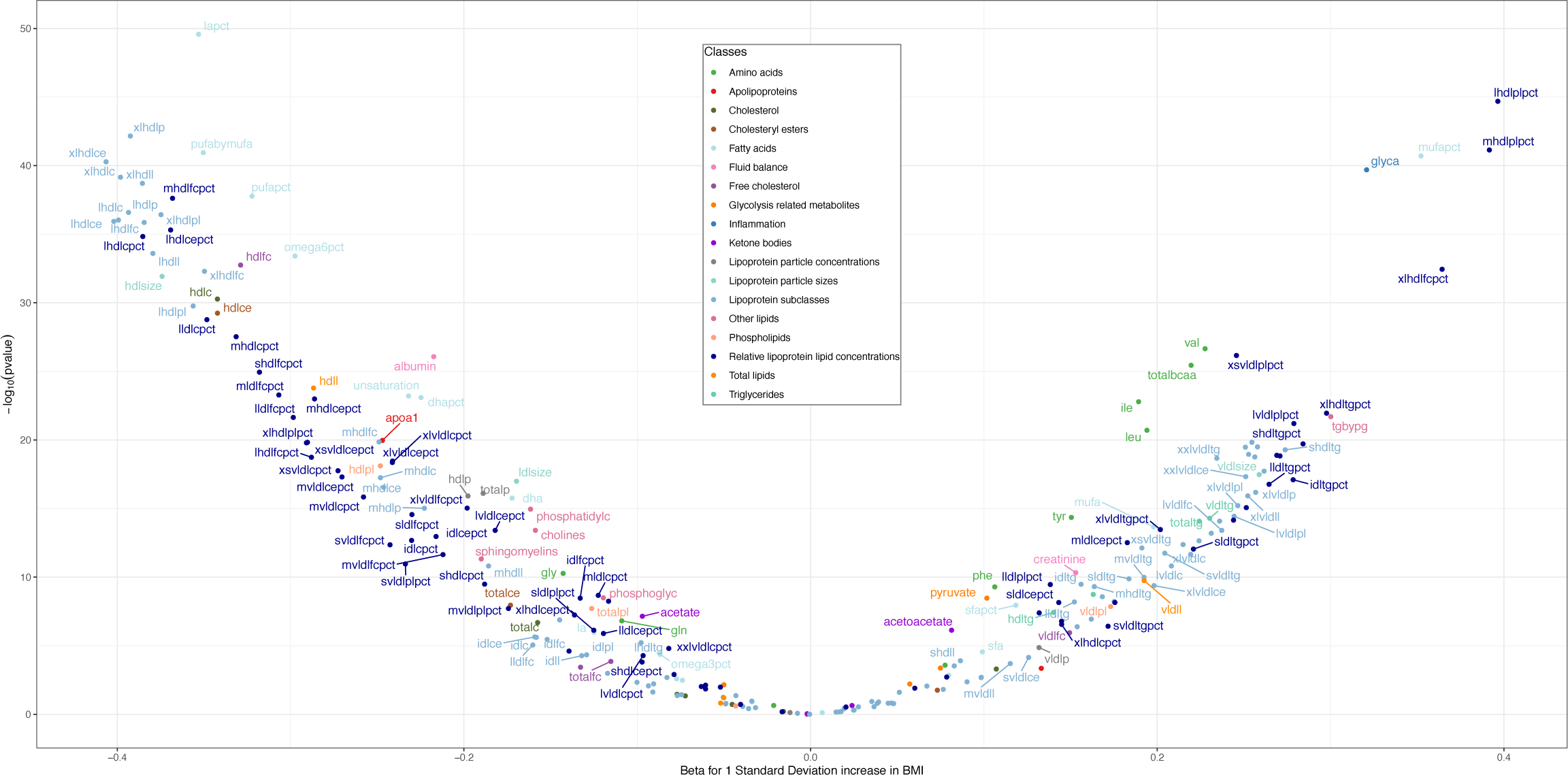
**Mendelian randomization estimates of the effect of BMI on metabolites** The volcano plot shows the estimates of the effect of BMI on circulating metabolites from Mendelian randomization analyses. Metabolites associated with BMI at *p*<0.05 after correcting for multiple testing using Bonferroni correction are labelled.

BMI had a broad effect on the metabolomic profile, with associations across many metabolite classes including numerous lipid metabolites and their ratios such as total cholesterol, total lipids, triglycerides and cholesteryl esters in high density lipoprotein (HDL) and very low-density lipoprotein (VLDL). For example, higher BMI led to 0.40 SD (95% CI = 0.34, 0.46) and 0.39 SD (95% CI = 0.38, 0.39) lower levels of total cholesterol in very large HDL in MR and observational analyses, respectively. There was also evidence that higher BMI increased levels of several amino acids including valine, isoleucine, leucine, tyrosine, and phenylalanine (**Supplementary Table 6**). For example, higher BMI led to 0.23 SD (95% CI = 0.19, 0.27) and 0.26 SD (95% CI = 0.25, 0.27) higher levels of valine in MR and observational analyses, respectively.

### Association between BMI-associated metabolites and endometrial cancer (Part III)

Of the 165 BMI-associated metabolites, 25 metabolites (representing 27 adiposity-metabolite-EC associations) were directionally consistent with an intermediate role in the association between BMI and overall EC (2 associations), endometrioid EC (1 association) and non-endometrioid EC (24 associations) from MR analyses (**Figure 4 and Supplementary Figure 3; Supplementary Table 8**). Observational analyses were only possible for overall EC. The two BMI-associated metabolites (phospholipid to total lipids ratio in medium VLDL and cholesteryl esters to total lipids ratio in medium HDL) had directions of effects consistent with MR analyses in the fully adjusted observational analysis (**Supplementary Table 9**).

**Figure 4.**
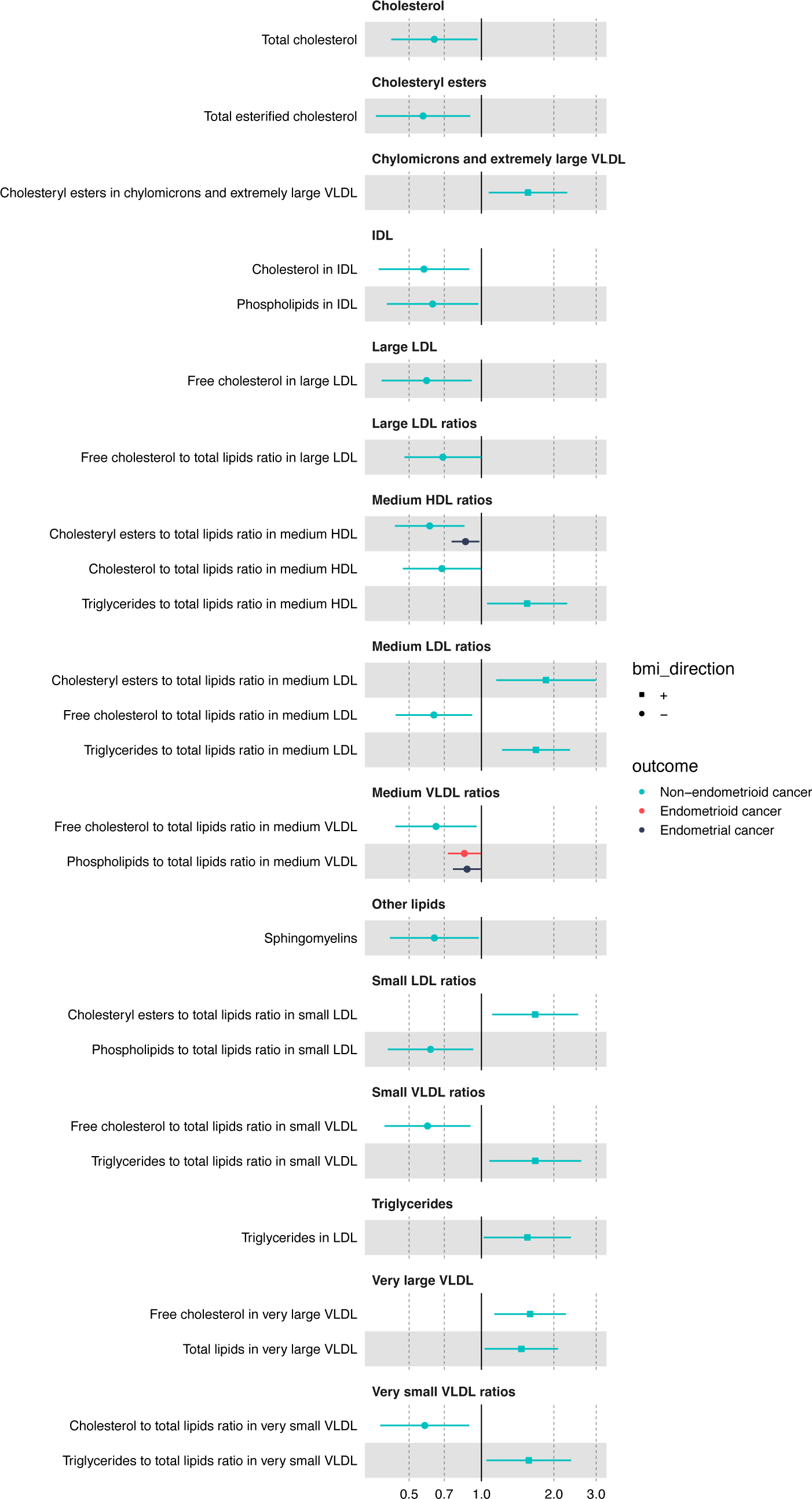
**Estimates of the causal effect of the BMI-associated metabolites on endometrial cancer risk** The forest plot shows the estimate of the causal effect of the BMI-associated metabolites and overall, endometrioid and non-endometrioid endometrial cancer based on two-sample MR (IVW method) from female-specific analyses. Only metabolites associated with endometrial cancer with a *p<*0.05 are shown in this figure. Symbols represent point estimates from individual analyses. Horizontal lines represent 95% confidence intervals. Squares represent metabolites that are positively associated with BMI and circles represent metabolites that are negatively associated with BMI.

Of the 25 BMI-associated metabolites, 2 were associated with two EC outcomes in the MR analyses: phospholipids to total lipids ratio in medium VLDL was associated with overall (OR = 0.87; 95% CI = 0.75, 1.00) and endometrioid EC (OR = 0.85, 95% CI = 0.72, 0.99) and cholesteryl esters to total lipids ratio in medium HDL was associated with overall (OR = 0.86, 95% CI = 0.75, 0.98) and non-endometrioid EC (OR = 0.61, 95% CI = 0.44, 0.85). The remaining 23 metabolites - mainly of the lipoprotein subclasses - were associated with just one EC outcome, which was predominantly non-endometrioid cancer. For example, the largest positive and negative effects were observed for the association between cholesteryl esters to total lipids ratio in medium LDL (OR = 1.85; 95% CI = 1.15, 2.98) and total esterified cholesterol (OR = 0.57; 95% CI = 0.36, 0.90) and non-endometrioid cancer using the IVW-MRE model (**Figure 4**).

### Examining the intermediate role of metabolites in the effect of adiposity on EC: Multivariable MR analyses (Part IV)

We used MVMR to investigate whether the 25 BMI-associated metabolites were potential intermediates of the effect of BMI on EC risk. In MVMR, there was little evidence that the association of BMI with overall and endometrioid EC was strongly attenuated following adjustment for most metabolites (**Figure 5)**. For non-endometrioid EC, the association of BMI with non-endometrial cancer was attenuated following adjustment for various metabolites (**Figure 5**); for example, the univariable MR OR for non-endometrioid EC per 1 SD higher BMI was 2.51 (95%CI = 1.47, 4.29), whereas the MVMR OR for non-endometrioid EC per 1 SD increase in BMI was 1.18 (95%CI = 0.53, 2.66) after adjusting for free cholesterol to total lipids ratio in medium LDL. The conditional F statistics for BMI and metabolites instruments are presented in **Supplementary Table 10**.

**Figure 5.**
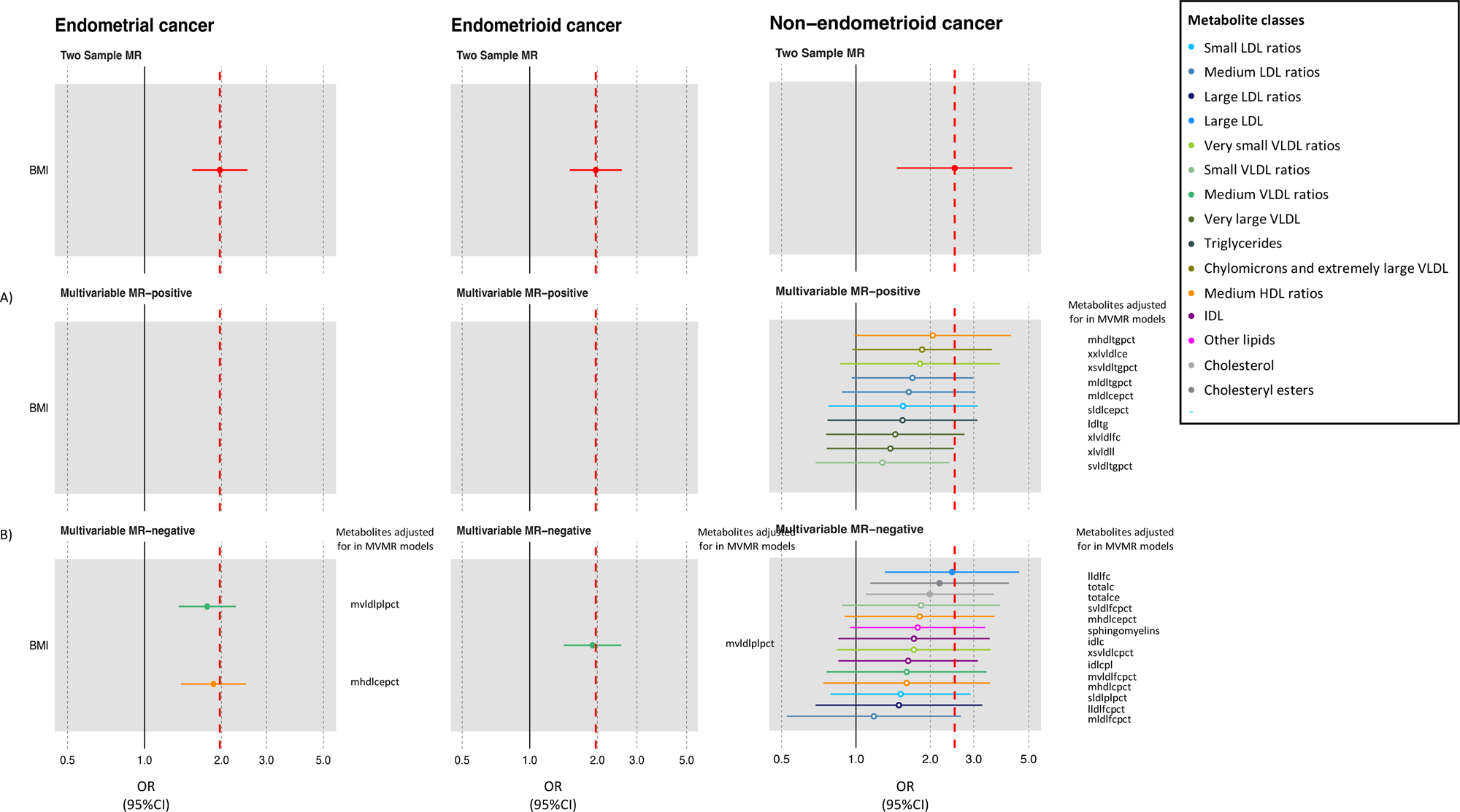
**Multivariable mendelian randomization estimates of the direct effect of BMI on endometrial cancer after adjustment for various BMI-associated metabolites** The forest plot shows the estimate of the direct causal effect of BMI on overall, endometrioid and non-endometrioid endometrial cancer based on multivariable MR analyses after adjustment for various metabolites that were associated with A) increased association with both BMI and endometrial cancer or B) decreased association with both BMI and endometrial cancer. Symbols represent point estimates of the direct effect of BMI on endometrial cancer after adjustment for various BMI-associated metabolites separately from multivariable MR analyses. Dotted red line represents the estimates of the indirect effect of BMI on endometrial cancer obtained from univariable IVW analyses.

#### Examining off-target effects

Given the highly pleiotropic nature of the metabolite instruments, it is possible that the instruments might be proxying other common, but unmeasured, biological pathways rather than having a direct effect through the metabolites. Using a commonly used catalogue of genetic association results (PhenoScanner^47,48^), we conducted a phenome scan for the genetic instruments proxying metabolites potentially underlying the adiposity-EC relationship and compared the traits association profiles with those of randomly selected metabolites. We identified a larger than expected proportion of instruments associated with several lipid traits including HDL, LDL, triglycerides and total cholesterol, as well as anthropometric (height and adiposity) and blood cell traits for the metabolites underlying the adiposity-EC relationship compared to the randomly selected metabolites. This suggests that the metabolite instruments could be proxying for or are a response read-out of other potentially shared biological pathways (**Figure 6**).

**Figure 6.**
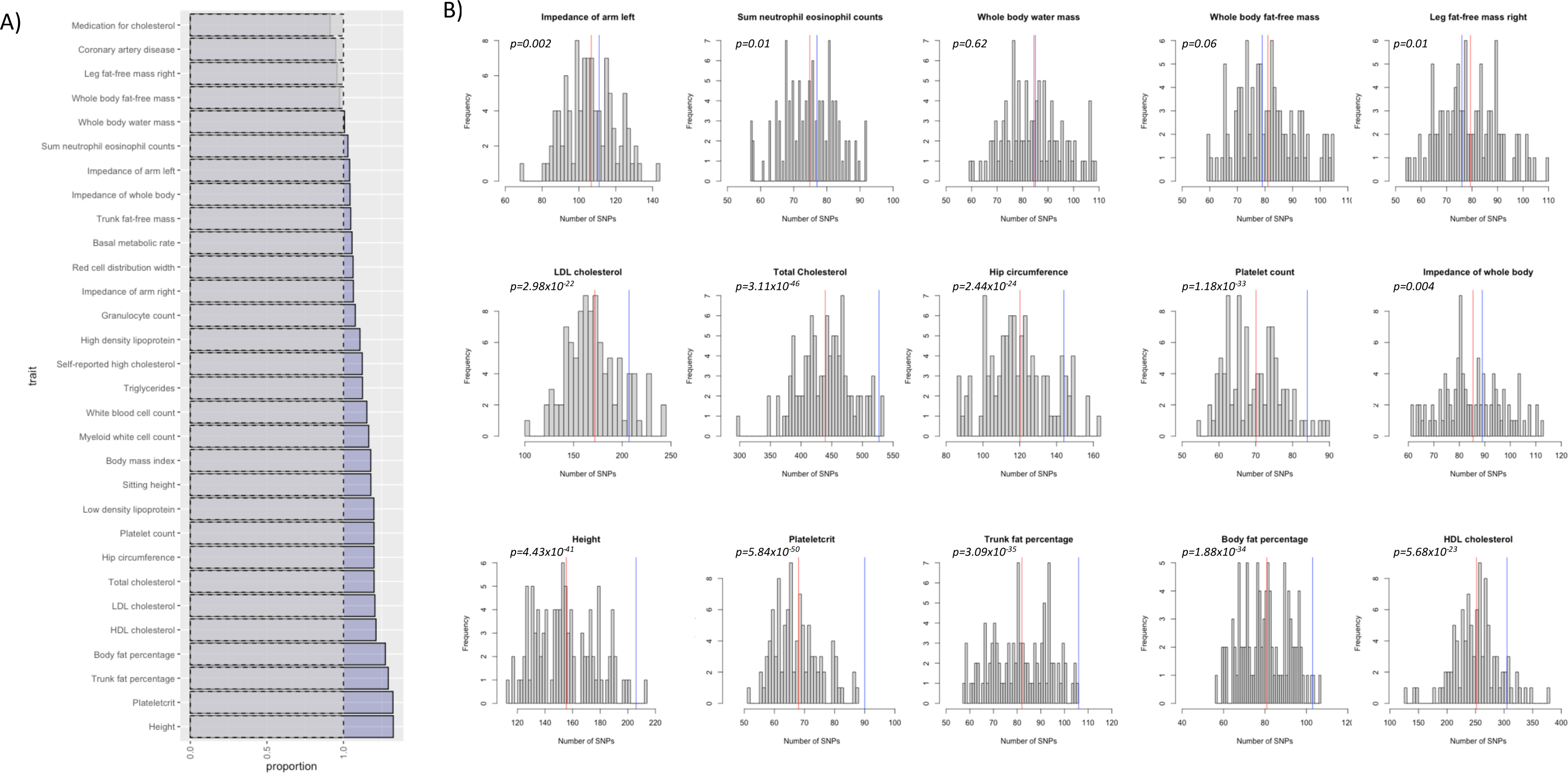
**Traits associated with genetic instruments for metabolites underlying effect of BMI on endometrial cancer** A) The blue bars represent the proportion of genetic instruments associated with the top 30 traits identified in PhenoScanner for metabolites underlying the effect of BMI on endometrial cancer relative to the negative control (grey bars). The negative control was obtained by iteratively (N=100) looking up traits associated with instruments for 25 randomly selected metabolites not underlying the association between BMI and endometrial cancer; B) the histograms represent the frequency of the number of SNPs associated with trait from the negative control analysis. The red line represents the mean of the number of SNPs associated with each trait for the negative control and the blue line represents the number of SNPs associated with the metabolites underlying the effect of BMI on endometrial cancer risk associated with each trait.

## Discussion

This study aimed to identify potential metabolic intermediates of the effects of adiposity on EC risk using MR and observational analyses. Higher BMI increased risk of overall EC and EC stratified by histological subtypes. BMI was associated with numerous metabolites; 25 of which were also found to increase EC risk. There was evidence from both two-step and multivariable MR analyses that the effect of BMI on EC was mediated strongly by several lipid metabolites; however, further bioinformatic analysis highlighted numerous potential pleiotropic and shared mechanisms that could potentially explain associations and patterns of intermediate effect.

Adiposity is a well-recognised risk factor for EC^49–51^. Our analyses found that higher BMI and WHR were positively associated with EC. In contrast, in MR analyses, we only found strong evidence that higher BMI increased EC risk. This is consistent with findings from recent MR studies where genetically elevated BMI, but not WHR, was found to be causally linked to EC^6,8,52,53^. It is well-established in the literature that obesity is associated with both endometrioid and non-endometrioid EC, with stronger associations of an effect on endometrioid EC, as compared to non-endometrioid EC^6,8,52,54–60^. Interestingly, the effect of BMI on non-endometrioid EC was slightly greater than endometrioid EC in our study. Although rarer, non-endometrioid tumours are more aggressive and have a poorer prognosis. Little is known about the etiology of non-endometrioid EC and the association with obesity warrants further investigation. WHR is widely used as a proxy measure of central adiposity and, notwithstanding variation in the statistical performance of analyses here and the impact of measurement error and the genetic architecture of these traits on the nature of MR analyses, these findings may suggest that central adiposity may be less relevant for EC risk than overall adiposity. This is supported by a recent MR study which found little evidence for a causal link between trunk fat ratio (proxy for abdominal adiposity) and EC but found strong evidence that BMI and arm fat ratio (proxy for overall body fat) had a strong positive effect on EC^53^.

Adiposity leads to perturbations in systemic metabolism which may have an impact on EC^16,61^. In both MR and observational analyses, BMI was associated with a wide array of metabolites; the effect estimates for a majority of metabolites (including HDL, VLDL, triglycerides, apolipoproteins A and B, glycolysis-related metabolites and amino acids) were generally consistent in terms of direction and magnitude with previous studies^15,41,62,63^; however, the associations of BMI with several metabolites including LDL cholesterol, total cholesterol, sphingomyelin and fatty acids were inconsistent with patterns previously observed in an MR study conducted in young adults^15^. For example, our study found evidence of an inverse effect of BMI on LDL cholesterol which is in contrast to the previous MR study^15^ which supported a positive effect of BMI. It has been suggested that the inconsistencies could be partly attributable to higher prevalence of statin use in UK Biobank (22% of men and 12% of women)^62^.

Of the BMI-associated metabolites, 25 metabolites were also associated with either overall, endometrioid or non-endometrioid EC. Our two-step and multivariable MR suggested that several lipid metabolites - the majority of which were metabolite ratios - could be potential intermediates between BMI and non-endometrioid EC risk. Cholesterol is an essential structural component of cell membranes and altered cholesterol metabolism can drive the onset and progression of cancer^64^. In the circulation and within cells, free cholesterol is converted to cholesteryl esters (CE) by acylCoA-cholesteryl-acyl-transferase (ACAT). Numerous studies have shown that increased expression and activity of ACAT and subsequent accumulation of cholesteryl esters in cancer cells is linked to the development and progression of cancers, including endometrial cancer^65,66^. We did not find strong evidence that total cholesterol esters attenuated the effect of BMI on non-endometrioid EC risk in our MVMR analyses. Interestingly, we found evidence that cholesteryl esters to total lipids ratio in small and medium LDL were positively associated with both BMI and non-endometrioid EC risk while free cholesterol to total lipids ratio in medium and large LDL were inversely associated with BMI and in turn associated with reduced risk of non-endometrioid EC. These metabolite ratios could represent flux through a biochemical pathway^67^ and further studies could be conducted to investigate their role in the development of EC.

Triglycerides, a major component of adipose tissues, are often elevated in adiposity and have been suggested to drive EC development either directly or indirectly through deregulation of peroxisome proliferator-activated receptors (PPARs)^68^. We observed that triglycerides in LDL and triglycerides to total lipids ratio in VLDL, LDL and LDL particles were positively associated with both BMI and non-endometrioid endometrial cancer risk. The positive relationship between triglycerides and endometrial cancer risk is consistent with several observational studies^68–71^; however, a recent Mendelian randomization analysis study found little evidence of a causal link between the role of triglycerides in the development of both endometrioid and non-endometrioid EC^72^. To this end, further research is required to clarify the role of triglycerides in non-endometrioid EC development.

Understanding how metabolites may be potential intermediates between risk factors and disease outcomes is a compelling research topic. However, major challenges persist for identifying instruments to proxy circulating metabolites due to the complex genetic architecture of blood metabolites ^44–46^. First, owing to the high correlational structure of many metabolites, instruments for metabolites are often associated with other metabolites^46,73^. Second, due to the high degree of pleiotropy (or basic biological overlap) for metabolite instruments with other modifiable risk factors and disease endpoints, it is often not straightforward to determine the exact molecular mechanism by which they impact the outcome. Indeed, when considering whether we are reliably instrumenting the effect of BMI-associated metabolites on EC, we identified other shared pathways including height and several adiposity and blood cell traits that could influence associations with EC. This suggests that when compared with a (hypothetically null) set of randomly selected metabolites, instruments for implicated metabolites could be flagging biological pathways that may be driving causal associations; however, importantly, it is difficult to distinguish whether the biological pathways highlighted are due to vertical pleiotropy (i.e., the mechanism by which the SNP influences the outcome is via metabolites) or horizontal pleiotropy (i.e., the SNP could influence the outcome through a pathway independent of metabolites). Our approach to the examination and characterisation of signal metabolites is conceptually similar to a cis-versus-trans MR analysis of proteins that looks to clarify inference from MR methods through prior information available on the instruments in question. Using this approach, a recent study^74^ demonstrated that the observed relationship between small HDL particle count and sepsis were - in part - driven by potential confounding between interleukin 6 (IL-6) and HDL, where IL-6 signalling is the true mechanism.

This work adds to the body of evidence suggesting a causal relationship between adiposity and adiposity-associated metabolites with overall, endometrioid, and non-endometrioid EC risk. In our study, we focussed predominantly on lipid-based metabolites identified via ^1^H-NMR metabolomics rather than mass spectrometry, which is not yet available at sufficient scale. Mass spectrometry-based metabolomic analyses offers a broader representation of metabolites beyond lipid subclasses and there is a growing body of evidence that metabolites detected by mass spectrometry are altered in EC patients^12,75^. Future metabolomic studies using mass spectrometry should be conducted to comprehensively evaluate the role of metabolites as intermediates between adiposity and EC.

Given the common shared genetic architecture, high correlation structure, and shared biology of metabolites, it is likely that a perturbation in any one metabolite does not happen in isolation. This is exemplified in recent work which has shown that the variance explained by a metabolite’s instrument is often greater for another^45,73,76^. Here, we use a naïve approach to instrumentation in a hypothesis generating analysis; however, given many metabolite instruments include only a handful of SNPs, statistical methods aiming to measure the potential effects of pleiotropy (e.g., MR-Egger) do not always lead to meaningful results. This also applies to techniques designed to evaluate the effect of multiple correlated exposures (e.g., MVMR).

In two-sample univariable MR analyses, sample overlap can bias estimates towards the confounded observational estimate (inflated type 1 error) in the presence of weak instruments in a manner proportional to the degree of overlap^42^. For our main analyses, we likely have 100% overlap between the adiposity and metabolites GWAS and up to a maximum 5% overlap for both of these with the overall EC GWAS. However, given bias due to sample overlap is negligible in the presence of strong instruments^77^ and that the results across sensitivity analyses using non-overlapping samples were generally consistent with the main analyses, it is unlikely that estimates from our analyses are meaningfully impacted by such bias.

Another limitation is the unrepresentative nature of UK Biobank (initial response rate ∼5%)^78^ and the potential for selection bias. Given the age of the UK Biobank participants, the prevalence of statin use is high (∼16% in UK Biobank vs ∼11% in the general UK adult population in 2014^79^). For our MR analyses, we used summary-level data from the ^1^H-NMR metabolite GWAS conducted using data from UK Biobank. This approach limits the capacity to fully explore the effects of other factors such as age and medication use (including statin use and HRT), which may influence the association between adiposity traits, metabolites and EC. Our analyses were limited to women of European ancestries; thus, these findings may not apply to individuals of other ancestries. Furthermore, given that we used the same ^1^H-NMR data from UK Biobank for both our MR and observational analyses examining the associations between adiposity traits and metabolites, these analyses cannot be considered independent. Replication of this study in other large cohort studies and in other ethnicities will allow a more robust characterisation of the metabolic profile associated with adiposity and the subsequent impact on EC.

## Conclusions

Our study suggests that higher BMI causes a higher risk of overall and all histological subtypes of EC and variation in numerous circulating metabolites. Several of these metabolites showed relationships consistent with an intermediate role in the effect of BMI on non-endometrioid EC from two-step and MVMR MR analyses; however, further bioinformatic analyses highlighted other potential shared mechanisms that could influence the risk of EC.

## Declarations

DP receives consulting fees from Johnson & Johnson and Novo Nordisk and payments for lectures, presentations, and educational events from Johnson & Johnson, Medtronic, and Novo Nordisk.

## Ethics approval

### Author contributions

MAL: conceptualization, data curation, formal analysis, investigation, methodology, project administration, validation, visualization, writing (original draft preparation), writing (review & editing)

VT: conceptualization, data curation, formal analysis, investigation, methodology, project administration, validation, visualization, writing (original draft preparation), writing (review & editing)

DP: resources, supervision, writing (review & editing)

SW: writing (review & editing)

LD: writing (review & editing)

MJG: writing (review & editing)

LJC: resources, supervision, writing (review & editing)

KHW: resources, supervision, writing (review & editing)

NJT: conceptualization, resources, supervision, writing (review & editing)

## Data Availability

The full summary statistics for BMI and WHR are available from the GIANT consortium (https://portals.broadinstitute.org/collaboration/giant/index.php/GIANT_consortium_data_files) and Zenodo (https://zenodo.org/record/1251813#.Yk7O25PMIUE), data files 4, 5, 7, and 8. The full summary statistics for the endometrial cancer GWAS are available from the OpenGWAS database (https://gwas.mrcieu.ac.uk/datasets/); IDs for endometrial cancer GWAS: ebi-a-GCST006464, ebi-a-GCST006465, and ebi-a-GCST006466; This can be accessed via the TwoSampleMR (https://mrcieu.github.io/TwoSampleMR/) and ieugwasr (http://gwas-api.mrcieu.ac.uk/) R packages or directly from OpenGWAS in GWAS-VCF format50. The full summary statistics for the 1H-NMR metabolites will be made available at the University of Bristol data repository. The individual level data used in this work is not publicly available and can only be obtained from UK Biobank with an approved application.

## Acknowledgments

We would like to express our appreciation to UK Biobank participants and staff for providing the research community with this valuable source of data. This work used data from application number 16391.

## Code and data availability

Univariable MR analyses were performed using TwoSampleMR^33^(version 0.4.22).; multivariable MR analyses were performed using MVMR)^39^ (version 0.3) R package; forest plots were created using ggforestplot (version 0.1)^80^ R package; circos plots were created using EpiViz^81–83^ (version 0.1) R package. All publicly available data, code, and results used in this work are available on GitHub: https://github.com/mattlee821/adiposity_metabolites_endometrial_cancer. This includes all exposure data used in all MR analyses. The full summary statistics for BMI and WHR are available from the GIANT consortium (https://portals.broadinstitute.org/collaboration/giant/index.php/GIANT_consortium_data_files) and Zenodo (https://zenodo.org/record/1251813#.Yk7O25PMIUE), data files 4, 5, 7, and 8. The full summary statistics for the endometrial cancer GWAS are available from the OpenGWAS database (https://gwas.mrcieu.ac.uk/datasets/); IDs for endometrial cancer GWAS: ebi-a-GCST006464, ebi-a-GCST006465, and ebi-a-GCST006466; This can be accessed via the TwoSampleMR (https://mrcieu.github.io/TwoSampleMR/) and ieugwasr (http://gwas-api.mrcieu.ac.uk/) R packages or directly from OpenGWAS in GWAS-VCF format^50^. The full summary statistics for the ^1^H-NMR metabolites will be made available at the University of Bristol data repository. The individual level data used in this work is not publicly available and can only be obtained from UK Biobank with an approved application.

## Funding

This work was supported by the Wellcome Trust through a Wellcome Trust Investigator award to NJT (202802/Z/16/Z, 2016-2023). MAL is funded by a Medical Research Council GW4 studentship (grant number: MR/R502340/1). NJT was a Wellcome Trust Investigator (202802/Z/16/Z, 2016-2023), is the PI of the Avon Longitudinal Study of Parents and Children (MRC & WT 217065/Z/19/Z), is supported by the University of Bristol NIHR Biomedical Research Centre (BRC-1215-2001), the MRC Integrative Epidemiology Unit (MC_UU_00011/1) and works within the CRUK Integrative Cancer Epidemiology Programme (C18281/A29019). VT, NJT, SW, LD and MG are supported by the Cancer Research UK (grant number: PRCPJT-May22\100028). L.J.C. was supported by N.J.T.’s Wellcome Investigator Award (202802/Z/16/Z) 2016-2023, and by MC_UU_00032/1 since 2023. All authors work in the MRC Integrative Epidemiology Unit at the University of Bristol, which is supported by the Medical Research Council (grant numbers: MC_UU_00011/1-7) and the University of Bristol. KHW is supported by the University of Bristol and Cancer Research UK [grant number RCCPDF\100007]. DJP has been funded by the Royal College of Surgeons of England. He receives consulting fees from Johnson & Johnson and Novo Nordisk and payments for lectures, presentations, and educational events from Johnson & Johnson, Medtronic, and Novo Nordisk. This research was funded in whole, or in part, by the Wellcome Trust [202802/Z/16/Z, 217065/Z/19/Z]. For the purpose of Open Access, the author has applied a CC BY public copyright licence to any Author Accepted Manuscript version arising from this submission.

## Disclaimer

Where authors are identified as personnel of the International Agency for Research on Cancer/World Health Organization, the authors alone are responsible for the views expressed in this article and they do not necessarily represent the decisions, policy or views of the International Agency for Research on Cancer/World Health Organization.

## Conflict of interest

None declared

## Notes

### Competing Interest Statement

DJP receives consulting fees from Johnson & Johnson and Novo Nordisk and payments for lectures, presentations, and educational events from Johnson & Johnson, Medtronic, and Novo Nordisk.

### Author Declarations

Ethical approval was obtained from the Northwest Multi-centre Research Ethics Committee (11/NW/0382).

