## Supplementary Table 1 for "Adiposity, metabolites and endometrial cancer risk: Mendelian randomization and Observational analyses"

| **Table S1. STROBE Statement—Checklist of items that should be included in reports of *case-control studies*** | | |  |
| --- | --- | --- | --- |
|  | **Item No** | **Recommendation** | **Paragraph number** |
| **Title and abstract** | 1 | (*a*) Indicate the study’s design with a commonly used term in the title or the abstract | Title |
|  |  | (*b*) Provide in the abstract an informative and balanced summary of what was done and what was found | Abstract |
| **Introduction** | | | |
| Background/rationale | 2 | Explain the scientific background and rationale for the investigation being reported | Introduction, Paragraph 1-3 |
| Objectives | 3 | State specific objectives, including any prespecified hypotheses | Introduction, Paragraph 4 |
| **Methods** | | | |
| Study design | 4 | Present key elements of study design early in the paper | Methods, Section “Analytical strategy” |
| Setting | 5 | Describe the setting, locations, and relevant dates, including periods of recruitment, exposure, follow-up, and data collection | Methods, Section “Study population” under “Observational analyses (Part I-III)” |
| Participants | 6 | (*a*) Give the eligibility criteria, and the sources and methods of case ascertainment and control selection. Give the rationale for the choice of cases and controls | Methods, Section “Study population” under “Observational analyses (Part I-III) |
|  |  | (*b*) For matched studies, give matching criteria and the number of controls per case | Not applicable |
| Variables | 7 | Clearly define all outcomes, exposures, predictors, potential confounders, and effect modifiers. Give diagnostic criteria, if applicable | Methods, Section “Study population” under “Observational analyses (Part I-III) |
| Data sources/ measurement | 8 | For each variable of interest, give sources of data and details of methods of assessment (measurement). Describe comparability of assessment methods if there is more than one group | Methods, Section “Study population” under “Observational analyses (Part I-III) |
| Bias | 9 | Describe any efforts to address potential sources of bias | Methods, Section “Statistical analysis” under “Observational analyses (Part I-III) |
| Study size | 10 | Explain how the study size was arrived at | Methods, Section “Study population” under “Observational analyses (Part I-III)” |
| Statistical methods | 12 | (*a*) Describe all statistical methods, including those used to control for confounding | Methods, Section “Statistical analysis” under “Observational analyses (Part I-III) |
|  |  | (*b*) Describe any methods used to examine subgroups and interactions | Not applicable |
|  |  | (*c*) Explain how missing data were addressed | Analyses only included the complete data and those with missing data were excluded |
|  |  | (*d*) If applicable, explain how matching of cases and controls was addressed | Not applicable |
|  |  | (*e*) Describe any sensitivity analyses | Methods, Section “Statistical analysis” under “Observational analyses (Part I-III) |
| **Results** |  |  |  |
| Participants | 13 | (a) Report numbers of individuals at each stage of study—eg numbers potentially eligible, examined for eligibility, confirmed eligible, included in the study, completing follow-up, and analysed | Results, “Population and data overview” |
|  |  | (b) Give reasons for non-participation at each stage | Not applicable |
|  |  | (c) Consider use of a flow diagram | Not applicable |
| Descriptive data | 14 | (a) Give characteristics of study participants (eg demographic, clinical, social) and information on exposures and potential confounders | Table 1 |
|  |  | (b) Indicate number of participants with missing data for each variable of interest | Not applicable |
| Outcome data | 15 | Report numbers in each exposure category, or summary measures of exposure | Table 1 |
| **Discussion** |  |  |  |
| Key results | 18 | Summarise key results with reference to study objectives | Discussion, Paragraph 1 |
| Limitations | 19 | Discuss limitations of the study, taking into account sources of potential bias or imprecision. Discuss both direction and magnitude of any potential bias | Discussion, Paragraph 5-10 |
| Interpretation | 20 | Give a cautious overall interpretation of results considering objectives, limitations, multiplicity of analyses, results from similar studies, and other relevant evidence | Discussion, Paragraph 2, 3 and 4 |
| Generalisability | 21 | Discuss the generalisability (external validity) of the study results | Discussion, Section “Strengths and limitations” |
| **Other information** | | | |
| Funding | 22 | Give the source of funding and the role of the funders for the present study and, if applicable, for the original study on which the present article is based | Funding |
