## Supplementary Table 2 for "Adiposity, metabolites and endometrial cancer risk: Mendelian randomization and Observational analyses"

| **Table S2. STROBE-MR checklist** |  |
| --- | --- |
| **Item** | **Complete/location** |
| **1. Title and Abstract:** "Mendelian randomization" is named both in the title and the abstract | “Mendelian randomization” is named in the title and abstract. |
| **Introduction** |  |
| **2.Background:** Explain the scientific background and rationale for the reported study. Is causality between exposure and outcome plausible? Justify why MR is a helpful method to address the study question. | Introduction, Paragraph 1-3 |
| **3.Objectives:** State specific objectives clearly, including pre-specified causal hypotheses (if any). | Introduction, Paragraph 3 and 4 and Methods, Section “Analytical strategy” |
| 4. **Study design and data sources:** Present key elements of study design early in the paper. Consider including a table listing sources of data for all phases of the study. For each data source contributing to the analysis, describe the following: a) Describe the study design and the underlying population from which it was drawn. Describe also the setting, locations, and relevant dates, including periods of recruitment, exposure, follow-up, and data collection, if available. b) Give the eligibility criteria, and the sources and methods of selection of participants. c) Explain how the analyzed sample size was arrived at. d) Describe measurement, quality and selection of genetic variants. e) For each exposure, outcome and other relevant variables, describe methods of assessment and, in the case of diseases, the diagnostic criteria used. f) Provide details of ethics committee approval and participant informed consent, if relevant. | Available information about the GWAS studies and selection of genetic variants is provided in the Methods, Section “Genome-wide association study and Mendelian randomisation analyses (Part I-IV)”. Further information is provided in each of the original GWAS publications. |
| **5. Assumptions:** Explicitly state assumptions for the main analysis (e.g. relevance, exclusion, independence, homogeneity) as well assumptions for any additional or sensitivity analysis. | Methods, Section “Genome-wide association study and Mendelian randomisation analyses (Part I-IV)”. |
| 6. **Statistical methods main analysis** Describe statistical methods and statistics used. a) Describe how quantitative variables were handled in the analyses (i.e., scale, units, model). b) Describe the process for identifying genetic variants and weights to be included in the analyses (i.e, independence and model). Consider a flow diagram. c) Describe the MR estimator, e.g. two-stage least squares, Wald ratio, and related statistics. Detail the included covariates and, in case of two-sample MR, whether the same covariate set was used for adjustment in the two samples. d) Explain how missing data were addressed. e) If applicable, say how multiple testing was dealt with. | a, b, c) Methods, Section “Genome-wide association study and Mendelian randomisation analyses (Part I-IV)”. d, e) Not applicable to our study |
| **7. Assessment of assumptions:** Describe any methods used to assess the assumptions or justify their validity. | Methods, Section “Genome-wide association study and Mendelian randomisation analyses (Part I-IV)”. |
| **8.Sensitivity analyses:** Describe any sensitivity analyses or additional analyses performed. | Methods, Section “Genome-wide association study and Mendelian randomisation analyses (Part I-IV)” and “Sensitivity analyses” |
| 9. **Software and pre-registration** a) Name statistical software and package(s), including version and settings used. b) State whether the study protocol and details were pre-registered (as well as when and where). | a) Methods, Section “Genome-wide association study and Mendelian randomisation analyses (Part I-IV)”. b) Not applicable to our study |
| **Results** |  |
| 10. **Descriptive data** a) Report the numbers of individuals at each stage of included studies and reasons for exclusion. Consider use of a flow-diagram. b) Report summary statistics for phenotypic exposure(s), outcome(s) and other relevant variables (e.g. means, standard deviations, proportions). c) If the data sources include meta-analyses of previous studies, provide the number of studies, their reported ancestry, if available, and assessments of heterogeneity across these studies. Consider using a supplementary table for each data source. d) For two-sample Mendelian randomization: i. Provide information on the similarity of the genetic variant-exposure associations between the exposure and outcome samples. ii. Provide information on extent of sample overlap between the exposure and outcome data sources. | a) Methods, Section “Genome-wide association study and Mendelian randomisation analyses (Part I-IV)”.  b) Supplementary Table S3 c) Methods, Section “Genome-wide association study and Mendelian randomisation analyses (Part I-IV)”. d) Methods, Section “Genome-wide association study and Mendelian randomisation analyses (Part I-IV)”, “Sensitivity analyses” and discussion |
| 11. **Main results** a) Report the associations between genetic variant and exposure, and between genetic variant and outcome, preferably on an interpretable scale (e.g. comparing 25th and 75th percentile of allele count or genetic risk score, if individual-level data available). b) Report causal effect estimate between exposure and outcome, and the measures of uncertainty from the MR analysis. Use an intuitive scale, such as odds ratio, or relative risk, per standard deviation difference. c) If relevant, consider translating estimates of relative risk into absolute risk for a meaningful time-period. d) Consider any plots to visualize results (e.g. forest plot, scatterplot of associations between genetic variants and outcome versus between genetic variants and exposure). | Our results are given in terms of betas or odds ratios and confidence intervals throughout the results section.  We visualize results using forest plots in Figures 2,4,5 and volcano plots in Figures 3 and Supplementary Figure 2. |
| 12. **Assessment of assumptions** a) Assess the validity of the assumptions. b) Report any additional statistics (e.g., assessments of heterogeneity, such as I2, Q statistic). | We assessed the validity using sensitivity analyses such as MR-Egger , weighted median and weighted mode analyses, described in the “Association between adiposity measures and endometrial cancer (Part I)” , “Association between adiposity measures and metabolites (Part II)” and “Association between BMI-associated metabolites and endometrial cancer (Part III) of the results section and presented in Supplementary Table S5,S6 and S8. |
| 13. **Sensitivity and additional analyses** a) Use sensitivity analyses to assess the robustness of the main results to violations of the assumptions. b) Report results from other sensitivity analyses (e.g., replication study with different dataset, analyses of subgroups, validation of instrument(s), simulations, etc.). c) Report any assessment of direction of causality (e.g., bidirectional MR). d) When relevant, report and compare with estimates from non-MR analyses. e) Consider any additional plots to visualize results (e.g., leave-one-out analyses). | Results, “Association between adiposity measures and endometrial cancer (Part I)” , “Association between adiposity measures and metabolites (Part II)” and “Association between BMI-associated metabolites and endometrial cancer (Part III) and presented in Supplementary Table S5,S6 and S8. |
| **Discussion** |  |
| **14. Key results** | Discussion, Paragraph 1 |
| 15. **Limitations** Discuss limitations of the study, taking into account the validity of the MR assumptions, other sources of potential bias, and imprecision. Discuss both direction and magnitude of any potential bias, and any efforts to address them. | Discussion, Paragraph 5-10 |
| 16. **Interpretations** a) Give a cautious overall interpretation of results considering objectives and limitations. Compare with results from other relevant studies. b) Discuss underlying biological mechanisms that could be modelled by using the genetic variants to assess the relationship between the exposure and the outcome. c) Discuss whether the results have clinical or policy relevance, and whether interventions could have the same size effect. | a, b) Discussion, Paragraph 1-5 c) Not applicable to our study |
| **17.            Generalizability:** | Discussion, Paragraph 9 |
| **18.            Funding:** | Funding |
| **19.            Data and data sharing:** | Data availability statement |
| **20.            Conflicts of Interest:** | Conflict of interest statement |
