## Supplementary figures for "Adiposity, metabolites and endometrial cancer risk: Mendelian randomization and Observational analyses"

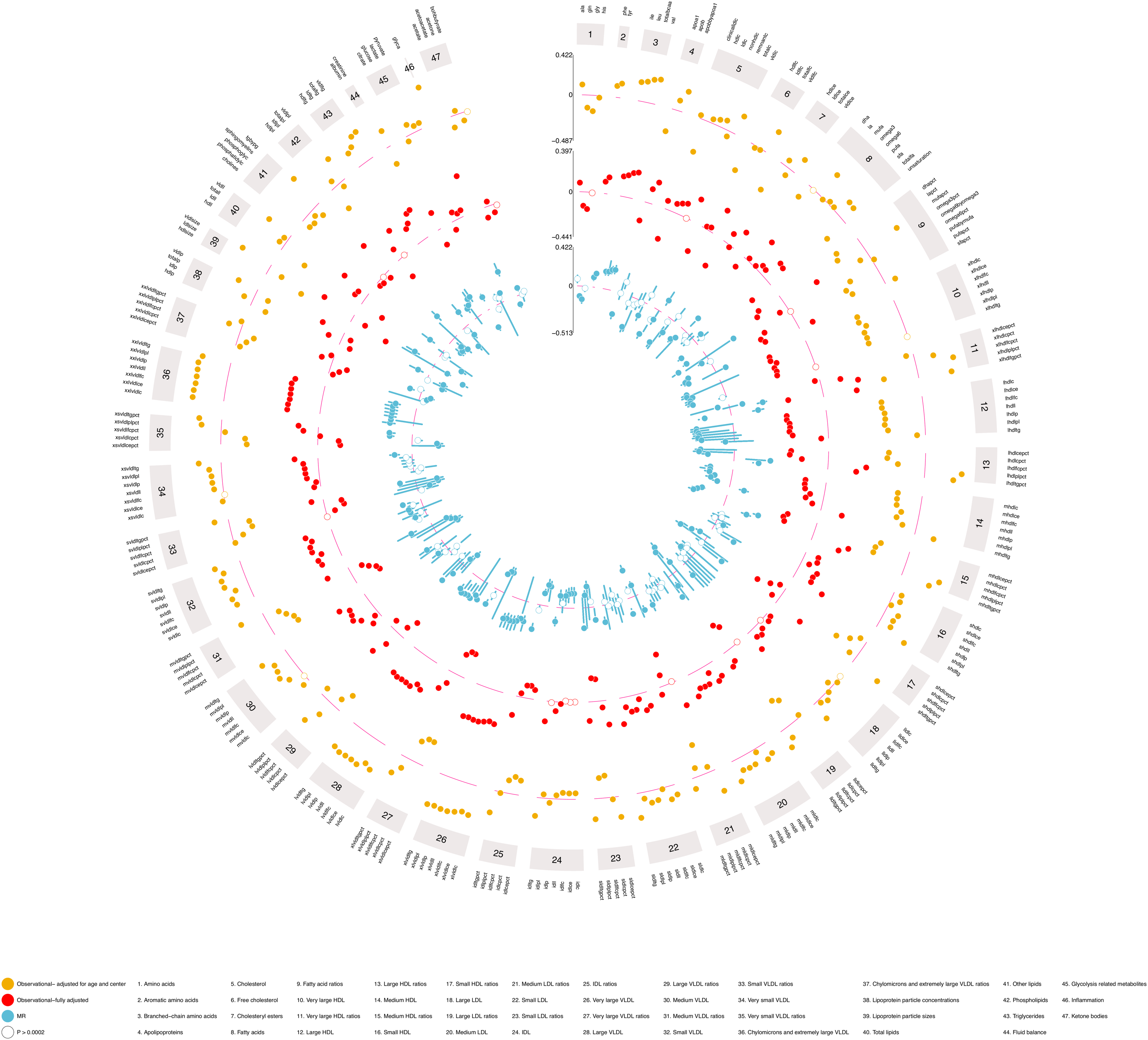

**Supplementary figure 1. MR estimates of the effect of BMI on metabolites**

The circos plot shows the estimates of the association between BMI and circulating metabolites from MR analyses (IVW method) using summary genome-wide association study (GWAS) data for BMI from the GIANT consortium (female-specific: N up to 434,794) and for the circulating metabolites from UK Biobank (female-specific: N up to 140,768). Symbols represent point estimates from individual analyses. Closed circles indicate *p*<0.05 after correcting for multiple testing using Bonferroni correction.

**Supplementary Figure 1. Observational estimates of the effect of BMI on metabolites**

The circos plot shows the estimates of the association between BMI and circulating metabolites from observational analyses in UK Biobank. Estimates from crude analyses (adjusted for age and centre) and fully adjusted analyses are represented in yellow and red, respectively. Estimates from MR estimates of the association between BMI and circulating metabolites are represented in blue. Symbols represent point estimates from individual analyses. Closed circles indicate *p*<0.05 after correcting for multiple testing using Bonferroni correction.

**Supplementary Figure 2. Comparison of Mendelian randomization and observational estimates of BMI and NMR metabolites**

A scatter plot of the MR estimates and the fully-adjusted observational estimates of association between BMI and NMR metabolites with a regression line (blue). Metabolites associated with BMI at *p*<0.05 after correcting for multiple testing using Bonferroni correction from MR analyses are labelled.

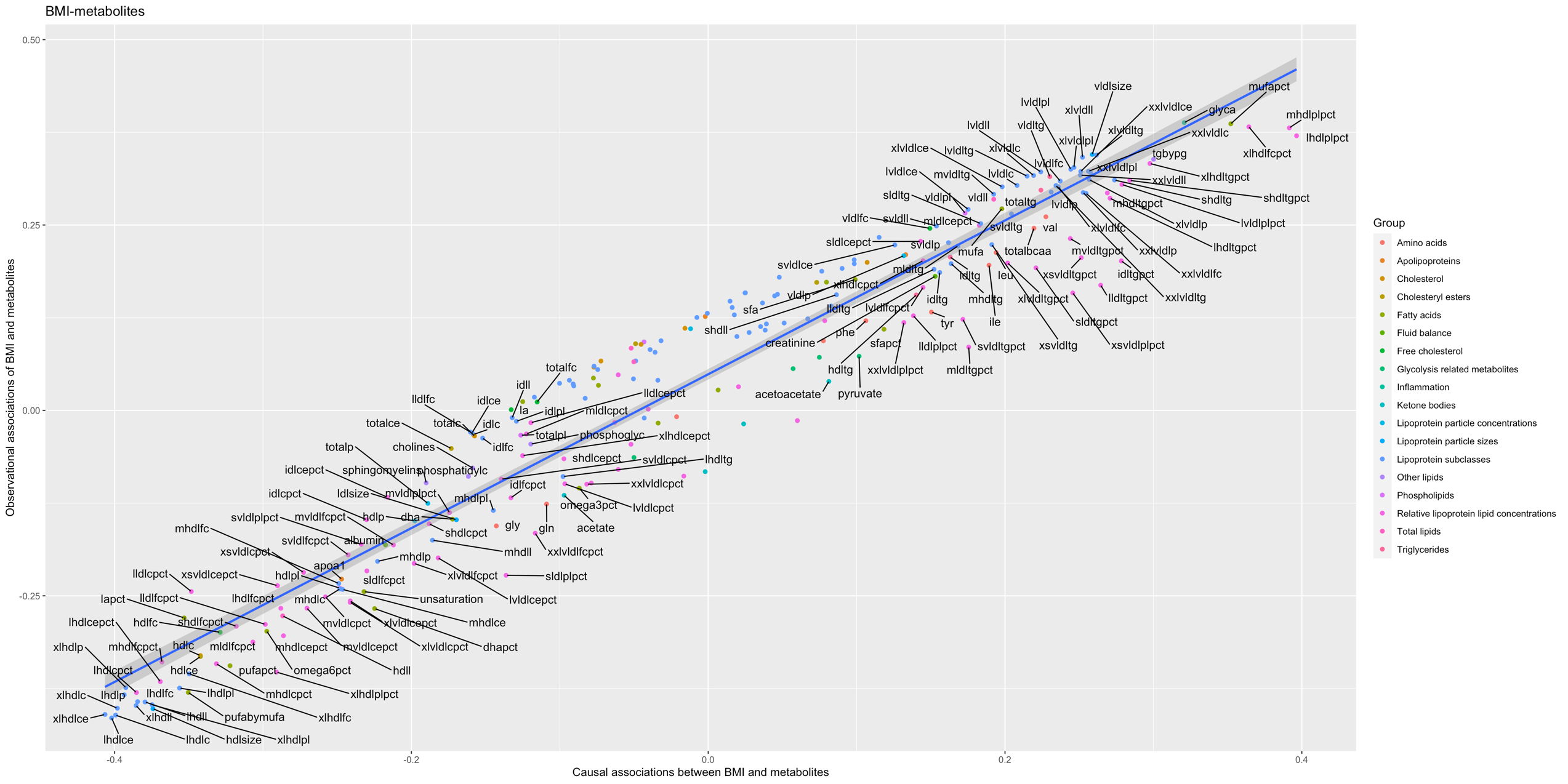

**r**=0.97

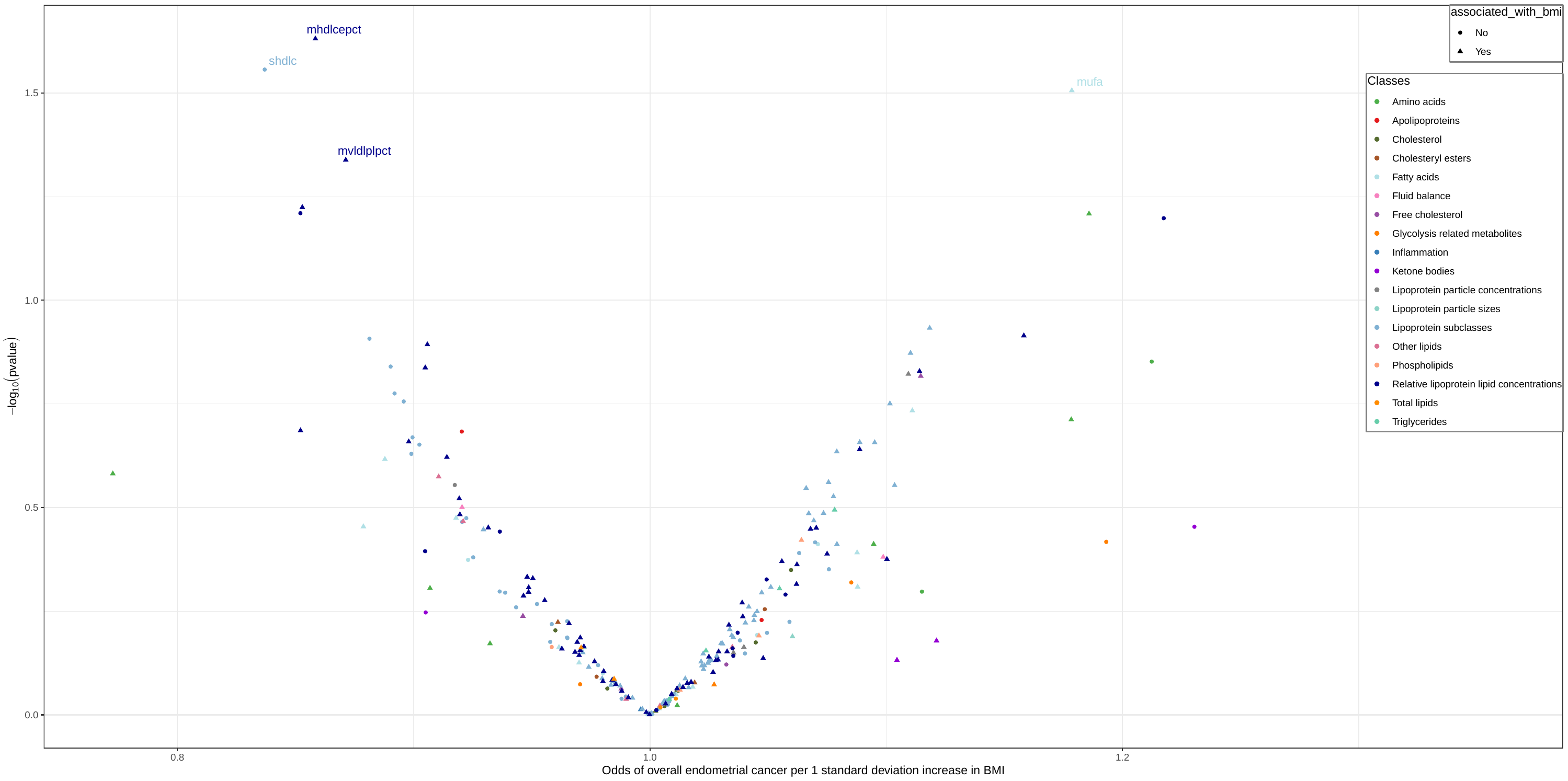

A)

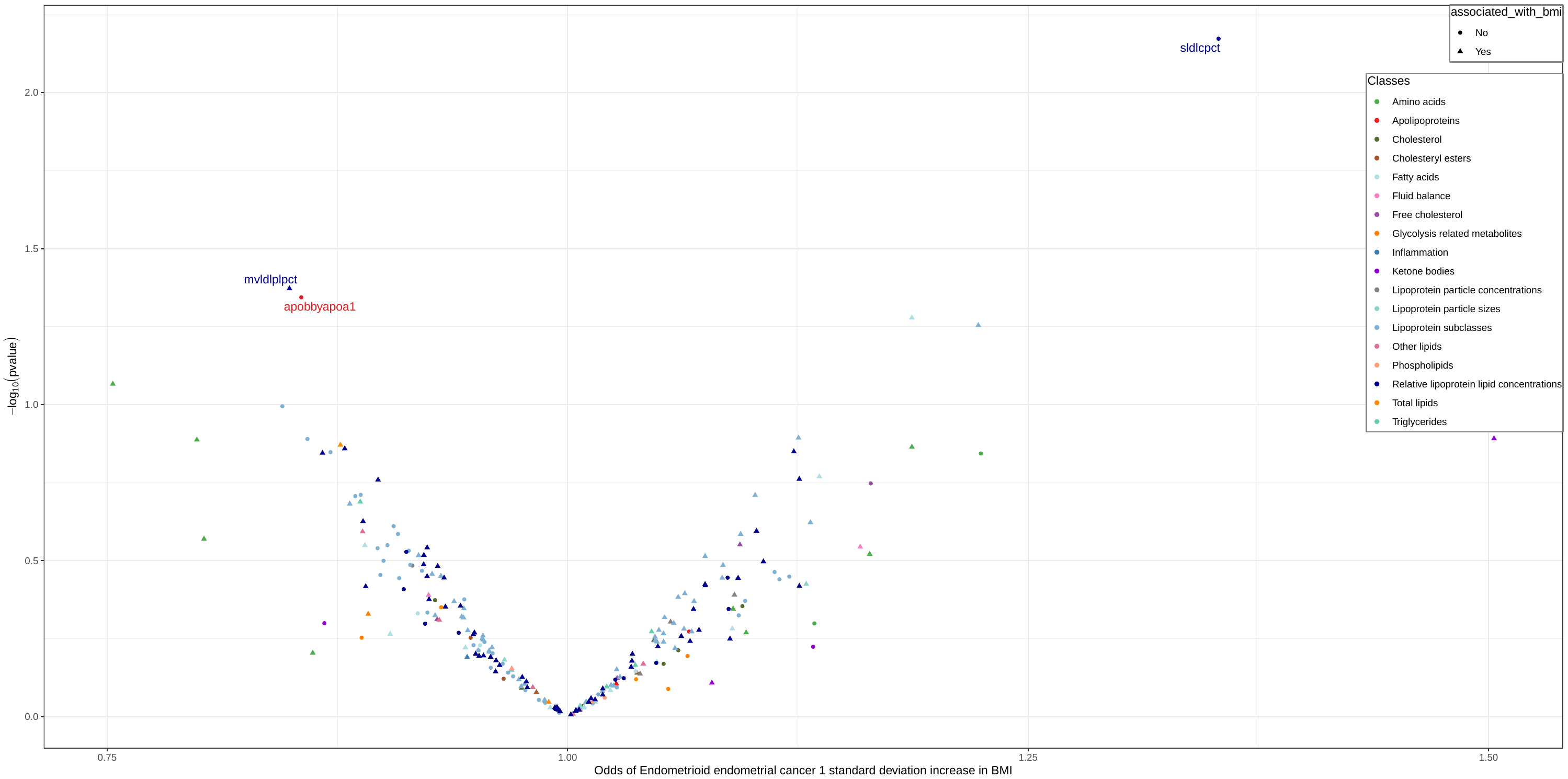

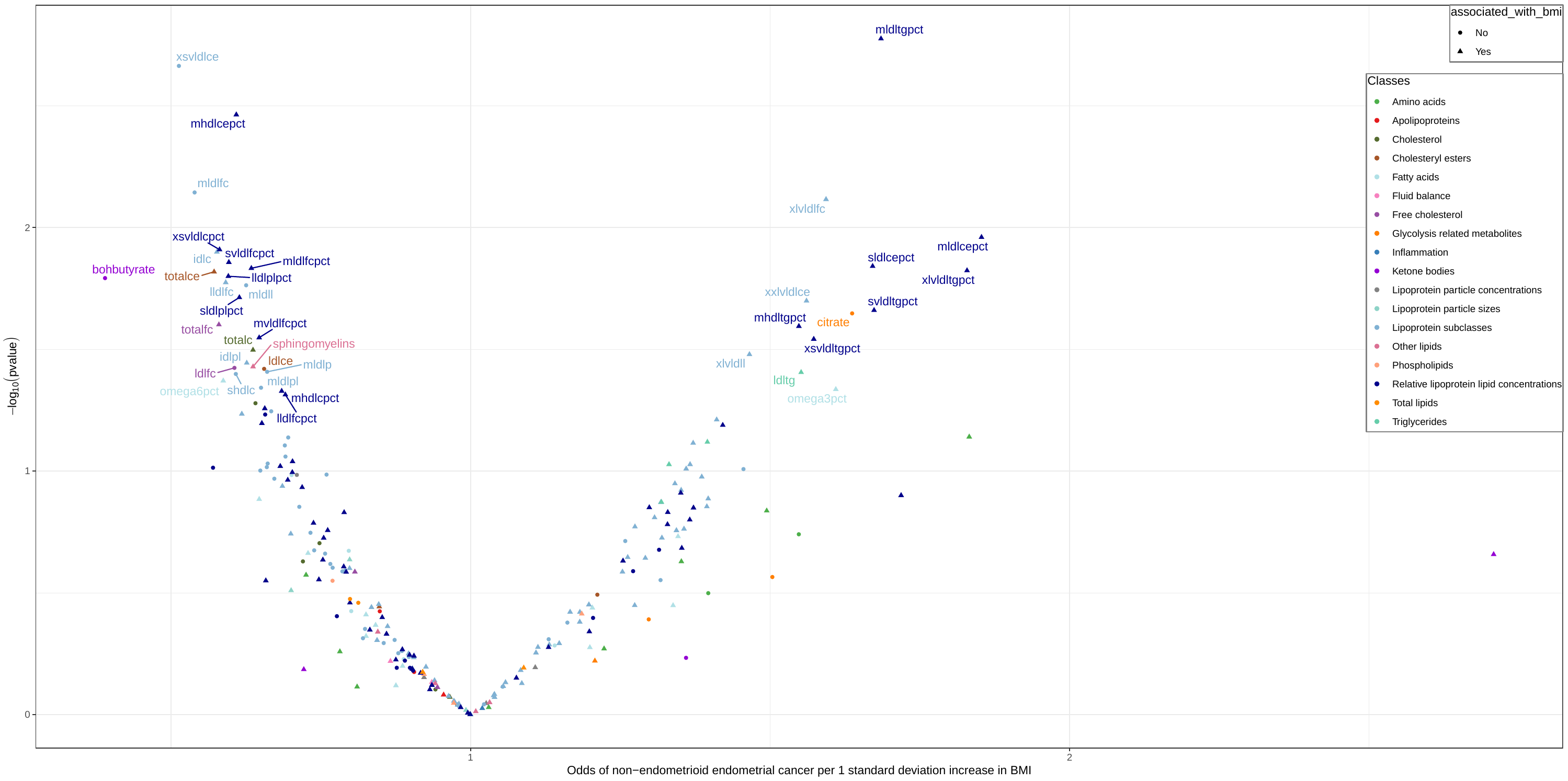

**Supplementary Figure 3. Mendelian randomization estimates of the effect of circulating metabolites on endometrial cancer.**

The volcano plot shows the estimates of the effect of circulating metabolites on A) overall endometrial cancer, B) endometrioid endometrial cancer and C) non-endometrioid endometrial cancer from Mendelian randomization analyses. Metabolites associated with endometrial cancer outcomes at *p*<0.05 after correcting for multiple testing using Bonferroni correction are labelled.

C)

B)
